# Cotton tipped plastic swabs for SARS-CoV2 RT-qPCR diagnosis to prevent supplies shortage

**DOI:** 10.1101/2020.04.28.20079947

**Authors:** Byron Freire-Paspuel, Patricio Vega-Mariño, Alberto Velez, Paulina Castillo, Ileana Elizabeth Gomez-Santos, Marilyn Cruz, Miguel Angel Garcia-Bereguiain

## Abstract

CDC and WHO guidelines for severe acute respiratory syndrome coronavirus 2 (SARS-CoV2) diagnosis only recommend synthetic fiber swabs for nasopharyngeal (NP) sampling. We show that cotton tipped plastic swabs do not inhibit PCR and have equivalent performance to rayon swabs. Cotton tipped plastic swabs are massively produced worldwide and would prevent swabs supplies shortage during current high SARS-CoV2 testing demands, particularly on developing countries.

## Introduction

NP swab is the reference sampling method for SARS CoV2 diagnosis, as recommended by World Health Organization (WHO) and Center for Diseases Control and Prevention (CDC) (1-3). CDC only endorses the use of synthetic fiber tipped swabs like rayon or nylon swabs on their recent guidelines for SARS-CoV2 diagnosis (3). WHO general guidelines for respiratory sample collection recommends either cotton or synthetic fiber swabs (2), but recent WHO guidelines for SARS-CoV2 diagnosis only endorse synthetic fiber swabs (1).

Multiple in vitro RT-qPCR diagnosis kits are available on the market for the detection of SARS-CoV2. Some of them have received emergency use authorization (EUA) from the U.S. Food & Drug Administration (FDA) while others only report validations made by manufacturers. The CDC designed 2019-nCoV CDC EUA kit (IDT, USA) is based on N1 and N2 probes to detect SARS-CoV-2 that have received positive evaluation on recent reports (3-5), and and RNase P as an RNA extraction quality control.

From the beginning of 2020, COVID 19 pandemia has rapidly widespread from Asia to Europe and USA, but also finally to Africa and Latin America. Public health systems have been challenged and definitely overflow in developing countries like Ecuador. In this context, the capacity to perform SARS-CoV2 tests is limited due to lack of enough laboratory equipment and trained personnel. Moreover, SARS-CoV2 diagnosis may be disrupted due to supplies shortage. For instance, Ecuador is experiencing supplies shortage of synthetic fiber swabs that is causing diagnosis disruption, particularly at isolated locations like Galapagos Islands where we implemented “LabGal” SARS-CoV2 diagnosis facility. Under this scenario, we conducted a validation study for NP sampling for SARS-CoV2 diagnosis using easily available cotton tipped plastic swabs with no inhibition effect over PCR reaction like wooden made ones.

## Methods

### Sample collection

A total of forty-four (44) subjects suspected of SARS-CoV2 infection during the surveillance implemented since April 7th 2020 at Galapagos Islands (Ecuador) were included on the study. All the subjects were tested for SARS-CoV2 using two different NP sterile plastic swabs: rayon tipped swabs and cotton tipped swabs (Puritan Medical Products LLC, USA; see supplementary image). Each NP swab was inserted in the nostril until they hit the back of the NP cavity, then rotated 5 times and removed. The test was conducted in both nostrils for each patient, with less than 2 minute of delay among each sample. NP swabs were immersed into a vial containing 0.5 mL TRIS-EDTA (pH 8) and keep refrigerated till arrival to the lab.

### Viral RNA extraction and RT-qPCR for SARS-CoV2

RNA extraction was performed using PureLink Viral RNA/DNA Mini Kit (Invitrogen, USA) following manufacturer’s instruction. Also, an extraction control (TRIS-EDTA pH 8) was done for each set of RNA extractions to exclude cross contamination.

SARS CoV2 was detected using RT-qPCR CDC protocol. Briefly, 2 different set of primers and probes (N1 and N2) are used for SARS-CoV2 detection while RNaseP primers and probe is the housekeeping product for RNA extraction quality control. Following CDC recommendations, the RT-qPCR kit selected was 2019-nCoV CDC EUA kit (IDT, USA). The assay was validated to detect 10 viral RNA copies/uL by using 2019-nCoV N positive control (IDT, USA) for N1 and N2 probes. All the experiments were performed using a CFX96 from BioRad.

### Statistics

For statistical analysis of Ct values, t-student test was performed using Excel.

### Ethics statement

The study was approved by “Comité de Operaciones de Emergencias Regional de Galápagos” that it is the board leading COVID19 surveillance at Galapagos Islands.

## Results

From the forty-four subjects included on the study, thirty-three (33; 75%) individuals were RT-qPCR SARS-CoV2 positive and eleven (11; 25%) were negative, either with plastic rayon tipped or plastic cotton tipped swabs (Table 1). Taking plastic rayon tipped swab NP sampling as the gold standard, the detection of SARS-CoV2 by plastic cotton tipped swab NP sampling yielded a 100% sensitivity and specificity, indicating a total agreement among swabs.

**Table 1.**
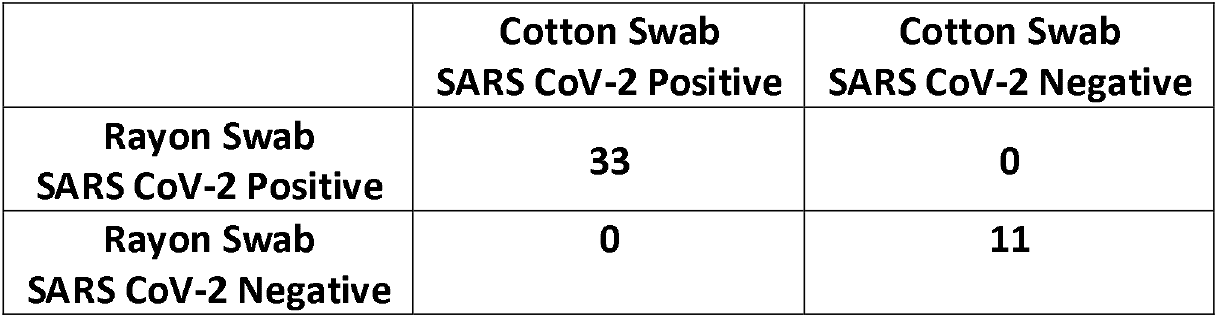
Plastic cotton tipped swabs and plastic rayon tipped swabs performance for NP sampling for SARS-CoV-2 RT-qPCR diagnosis.

|  | **Cotton Swab**<br>**SARS CoV-2 Positive** | **Cotton Swab**<br>**SARS CoV-2 Negative** |
| --- | --- | --- |
| **Rayon Swab**<br>**SARS CoV-2 Positive** | **33** | **0** |
| **Rayon Swab**<br>**SARS CoV-2 Negative** | **0** | **11** |

Ct (mean±SD) values for N1, N2 and RNaseP amplicons for plastic rayon tipped swabs (N1: 33.71±3.93; N2: 36.84±3.17; RNaseP: 33.75±3.05) and plastic cotton tipped swabs (N1: 32.55±5.14; N2: 34.37±5.25; RNaseP: 27.66±2.95) were statistically not different for viral specific amplicons N1 and N2 (p = 0.30 and 0.052, respectively), but statistically significant (p<0.001) for the RNA extraction quality control housekeeping gene RNaseP, indicating a better RNA extraction yield for plastic cotton tipped swabs (Table 2).

**Table 2.**
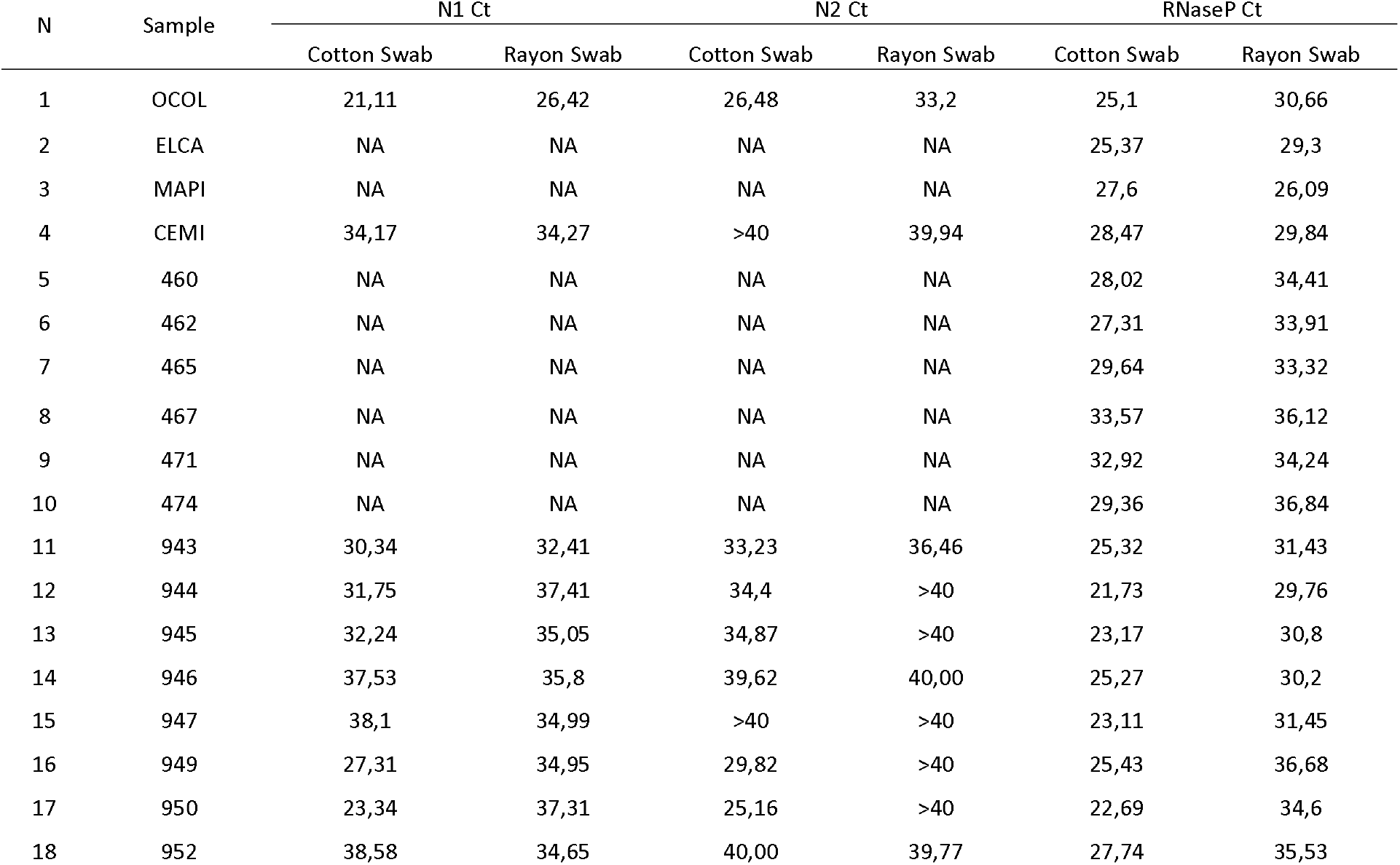
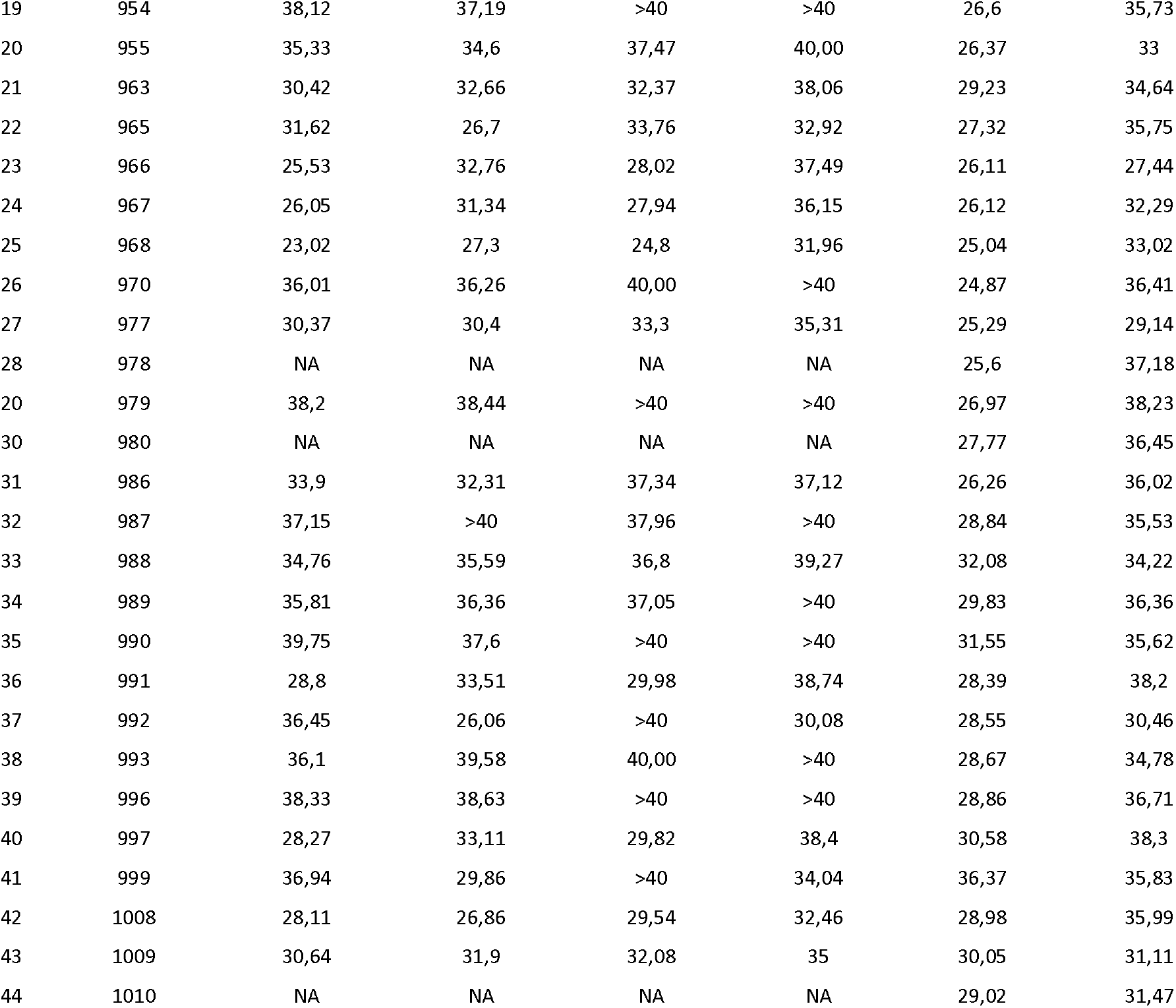
RT-qPCR Ct values for N1, N2 and RNaseP probes for nasopharyngeal samples with cotton and rayon swabs (mean +/- SD). NA mean “not amplified”.

| N | Sample | N1 Ct |  | N2 Ct |  | RNaseP Ct |  |
| --- | --- | --- | --- | --- | --- | --- | --- |
|  |  | Cotton Swab | Rayon Swab | Cotton Swab | Rayon Swab | Cotton Swab | Rayon Swab |
| 1 | OCOL | 21,11 | 26,42 | 26,48 | 33,2 | 25,1 | 30,66 |
| 2 | ELCA | NA | NA | NA | NA | 25,37 | 29,3 |
| 3 | MAPI | NA | NA | NA | NA | 27,6 | 26,09 |
| 4 | CEMI | 34,17 | 34,27 | >40 | 39,94 | 28,47 | 29,84 |
| 5 | 460 | NA | NA | NA | NA | 28,02 | 34,41 |
| 6 | 462 | NA | NA | NA | NA | 27,31 | 33,91 |
| 7 | 465 | NA | NA | NA | NA | 29,64 | 33,32 |
| 8 | 467 | NA | NA | NA | NA | 33,57 | 36,12 |
| 9 | 471 | NA | NA | NA | NA | 32,92 | 34,24 |
| 10 | 474 | NA | NA | NA | NA | 29,36 | 36,84 |
| 11 | 943 | 30,34 | 32,41 | 33,23 | 36,46 | 25,32 | 31,43 |
| 12 | 944 | 31,75 | 37,41 | 34,4 | >40 | 21,73 | 29,76 |
| 13 | 945 | 32,24 | 35,05 | 34,87 | >40 | 23,17 | 30,8 |
| 14 | 946 | 37,53 | 35,8 | 39,62 | 40,00 | 25,27 | 30,2 |
| 15 | 947 | 38,1 | 34,99 | >40 | >40 | 23,11 | 31,45 |
| 16 | 949 | 27,31 | 34,95 | 29,82 | >40 | 25,43 | 36,68 |
| 17 | 950 | 23,34 | 37,31 | 25,16 | >40 | 22,69 | 34,6 |
| 18 | 952 | 38,58 | 34,65 | 40,00 | 39,77 | 27,74 | 35,53 |
| 19 | 954 | 38,12 | 37,19 | >40 | >40 | 26,6 | 35,73 |
| 20 | 955 | 35,33 | 34,6 | 37,47 | 40,00 | 26,37 | 33 |
| 21 | 963 | 30,42 | 32,66 | 32,37 | 38,06 | 29,23 | 34,64 |
| 22 | 965 | 31,62 | 26,7 | 33,76 | 32,92 | 27,32 | 35,75 |
| 23 | 966 | 25,53 | 32,76 | 28,02 | 37,49 | 26,11 | 27,44 |
| 24 | 967 | 26,05 | 31,34 | 27,94 | 36,15 | 26,12 | 32,29 |
| 25 | 968 | 23,02 | 27,3 | 24,8 | 31,96 | 25,04 | 33,02 |
| 26 | 970 | 36,01 | 36,26 | 40,00 | >40 | 24,87 | 36,41 |
| 27 | 977 | 30,37 | 30,4 | 33,3 | 35,31 | 25,29 | 29,14 |
| 28 | 978 | NA | NA | NA | NA | 25,6 | 37,18 |
| 20 | 979 | 38,2 | 38,44 | >40 | >40 | 26,97 | 38,23 |
| 30 | 980 | NA | NA | NA | NA | 27,77 | 36,45 |
| 31 | 986 | 33,9 | 32,31 | 37,34 | 37,12 | 26,26 | 36,02 |
| 32 | 987 | 37,15 | >40 | 37,96 | >40 | 28,84 | 35,53 |
| 33 | 988 | 34,76 | 35,59 | 36,8 | 39,27 | 32,08 | 34,22 |
| 34 | 989 | 35,81 | 36,36 | 37,05 | >40 | 29,83 | 36,36 |
| 35 | 990 | 39,75 | 37,6 | >40 | >40 | 31,55 | 35,62 |
| 36 | 991 | 28,8 | 33,51 | 29,98 | 38,74 | 28,39 | 38,2 |
| 37 | 992 | 36,45 | 26,06 | >40 | 30,08 | 28,55 | 30,46 |
| 38 | 993 | 36,1 | 39,58 | 40,00 | >40 | 28,67 | 34,78 |
| 39 | 996 | 38,33 | 38,63 | >40 | >40 | 28,86 | 36,71 |
| 40 | 997 | 28,27 | 33,11 | 29,82 | 38,4 | 30,58 | 38,3 |
| 41 | 999 | 36,94 | 29,86 | >40 | 34,04 | 36,37 | 35,83 |
| 42 | 1008 | 28,11 | 26,86 | 29,54 | 32,46 | 28,98 | 35,99 |
| 43 | 1009 | 30,64 | 31,9 | 32,08 | 35 | 30,05 | 31,11 |
| 44 | 1010 | NA | NA | NA | NA | 29,02 | 31,47 |

## Discussion

We herein report that molecular detection of SARS-CoV2 using plastic cotton tipped swabs NP sampling is as reliable as using plastic synthetic fiber tipped like rayon swabs, considered as the gold standard by CDC (3). The main limitation of the study is the relative small sample size that would explain the 100% agreement among swabs. However, we believe that a potential disagreement among swabs on a bigger sample size study would be related to variability associated to sampling procedure more than to the type of swabs. While our results show that cotton does not inhibit the detection of SARS-COV2, previous work has shown inhibition by the chemicals in the wood stem of some swabs. This may explain why inexpensive cotton swabs have been excluded from CDC and WHO guidelines for SARS-CoV-2 diagnosis (1,3). However, the use of cotton tipped swabs for respiratory specimen collection is included at WHO general guidelines for respiratory specimen collection (2) and it has been reported specifically reliable for respiratory retroviruses like influenza (6).

Plastic cotton tipped swabs are cheap and made worldwide, even in developing countries like Ecuador. Including this type of swab on international guidelines upon more independent validation studies would help to prevent SARS-CoV2 diagnosis disruption due to swab supply shortage as recently happened in Ecuador, while keeping high standards for sensitivity and specificity.

To our knowledge, this is the the second study comparing swabs for SARS-CoV2 testing (7), but the first study suggesting that inexpensive, readily available cotton swabs could serve as a practical alternative to more costly, imported rayon swab. Additionally, high sensitivity was recently reported for nasal versus NP sampling for SARS-CoV2 diagnosis (8). Taking together this finding and ours, even sterile short plastic cotton tipped swabs like the one use for ear hygiene could represent an alternative under lack of NP swabs supply. We call upon the worldwide microbiology community, particularly at developing countries, to consider those findings and perform more validation studies to endorse plastic cotton swabs for SARS-CoV2 diagnosis to enhance the testing capacity to fight the spread of the current COVID-19 pandemic.

## Data Availability

Any data will be available upon request.

## Acknowledgments

We thank the medical personnel from “Ministerio de Salud Pública” at Galapagos Islands and the staff from the “Agencia de Regulación y Control de la Bioseguridad y Cuarentena para Galápagos” for their support. We also thank Dr. Ronald Cedeño from OPS/WHO for his work during Covid 19 surveillance in Galapagos Islands. We specially thank Gabriel Iturralde, Oscar Espinosa and Dr Tannya Lozada from “Dirección General de Investigación de la Universidad de Las Américas” for logistic support to make SARS-CoV2 diagnosis possible in Galapagos Islands.

## Disclaimers

The authors declare no conflict of interest.

**Supplementary image. Cotton and Rayon tipped plastic swabs used on the study (see the attached file)**.

